# REVEL is better at predicting pathogenicity of loss-of-function than gain-of-function variants

**DOI:** 10.1101/2023.06.06.23290963

**Authors:** Jasmin J Hopkins, Matthew N Wakeling, Matthew B Johnson, Sarah E Flanagan, Thomas W Laver

**Author notes:** **Corresponding author** Dr Thomas W Laver, Address: RILD Building, University of Exeter Medical School, Barrack Road, Exeter, EX2 5DW, UK. These authors contributed equally to this work (Sarah E Flanagan, Thomas W Laver).

## Abstract

*In silico* predictive tools can help determine the pathogenicity of variants. The 2015 American College of Medical Genetics and Genomics (ACMG) guidelines recommended that scores from these tools can be used as supporting evidence of pathogenicity. A subsequent publication by the ClinGen Sequence Variant Interpretation Working Group suggested high scores from some tools were sufficiently predictive to be used as moderate or strong evidence of pathogenicity.

REVEL is a widely used meta-predictor that uses the scores of 13 individual *in-silico* tools to calculate pathogenicity of missense variants. Its ability to predict missense pathogenicity has been assessed extensively, however, no study has previously tested whether its performance is affected by whether the missense variant acts via a loss of function (LoF) or gain of function (GoF) mechanism.

We used a highly curated dataset of 66 confirmed LoF and 65 confirmed GoF variants to evaluate whether this affected the performance of REVEL.

98% of LoF and 100% of GoF variants met the author-recommended REVEL threshold of 0.5 for pathogenicity, while 89% LoF and 88% GoF variants exceeded the 0.75 threshold. However, while 55% of LoF variants met the threshold recommended for a REVEL score to count as strong evidence of pathogenicity from the ACMG guidelines (0.932), only 35% of GoF variants met this threshold (P=0.0352).

GoF variants are therefore less likely to receive the highest REVEL scores which would enable the REVEL score to be used as strong evidence of pathogenicity. This has implications for classification with the ACMG guidelines as GoF variants are less likely to meet the criteria for pathogenicity.

## Introduction

*In silico* predictive tools can be used to help predict pathogenicity of genetic variants in Mendelian disease. They are particularly useful for missense variants since these have a variable effect on the protein: even in genes where missense variants are a known cause of disease, not all missense variants will be pathogenic.

As part of the standardisation of the classification of variants causing Mendelian disease the 2015 American College of Medical Genetics and Genomics (ACMG) guidelines stated that *in silico* predictive tools can be used as supporting evidence in variant classification ^1^ (PP3 criteria to support a variant being pathogenic and BP4 to support a variant being benign). The guidelines stratified the different lines of evidence that can be used to support a classification of pathogenic into different weights: supporting, moderate, strong, very strong. These different lines of weighted evidence are then combined to produce an overall variant classification of either benign, likely benign, uncertain significance, likely pathogenic or pathogenic. By classifying predictions from *in silico* tools as only supporting evidence they suggested limited weight could be put on their results. However, Pejaver *et al* ^2^, as part of the ClinGen ^3^ Sequence Variant Interpretation Working Group, recommended that some tools were sufficiently predictive of pathogenicity that high scores could be used as moderate (PP3_moderate) or even strong (PP3_strong) evidence for pathogenicity.

REVEL (Rare Exome Variant Ensemble Learner) is a meta-predictor - an *in silico* tool that combines multiple different tools and types of evidence for pathogenicity into a combined score ^4^. It uses scores from 13 individual tools: MutPred ^5^, FATHMM v2.3 ^6^, VEST 3.0 ^7^, PolyPhen-2 ^8^, SIFT ^9^, PROVEAN ^10^, MutationAssessor ^11^, MutationTaster ^12^, LRT ^13^, GERP++ ^14^, SiPhy ^15^, phyloP ^16^, and phastCons ^17^ to predict the likelihood that missense variants are pathogenic. This means that REVEL uses multiple strands of evidence to predict whether a variant is pathogenic: conservation, the difference in the physicochemical characteristics of the new amino acid compared to the reference and the effect of the amino acid change on the structural and functional properties of the protein.

REVEL is widely used in a range of applications and can have clinical implications. Toratani *et al*. ^18^ used REVEL to highlight a potential pathogenic variant in *RUNX1* predisposing to acute myeloid leukemia in a family, which led to choosing a stem cell donor from outside the family. Schuurmans e*t al*. ^19^ explored genotype-phenotype correlation in glutaric aciduria type 1 and showed that a higher REVEL score correlated with lower residual enzyme activity. Kingdom *et al*. ^20^ used REVEL to identify likely deleterious variants in genes associated with developmental disorders in order to screen the UK Biobank population cohort of 500,000 people for related phenotypes. As these examples highlight, REVEL is particularly useful as an automated assessment of pathogenicity, which can be used to take a cautious approach to pathogenicity (as in the transplant example). REVEL scores can also be easily correlated with other data, such as functional domains and enzymatic activity in the glutaric aciduria type 1 example. Finally, the scores offer the ability to classify a large number of variants in order to study the broad picture of a disease or phenotype in a large cohort where manual curation of variants may not be practical.

Gunning *et al*. ^21^ demonstrated that meta predictors, such as REVEL, provide superior predictive value over individual *in silico* tools. They analysed a dataset of variants from ClinVar, Human Gene Mutation Database (HGMD) and the Genome Aggregation Database (GnomAD) as well as a clinically representative dataset derived from research/diagnostic exome and panel sequencing. REVEL had the best performance of the meta-predictors tested on the results of the clinically representative dataset with an area under the receiver operating characteristic curve of 0.82. However, this study did not test whether the mechanism of action, loss of function (LoF) or gain of function (GoF), had an impact on REVEL’s performance.

REVEL produces a score for a missense variant of between 0 and 1 with larger scores indicating a higher chance that the variant is pathogenic. In the paper describing the tool, the authors give two potential thresholds for considering a variant to be pathogenic: a REVEL score of 0.5, which in their dataset (a subset of HGMD) gave a sensitivity of 0.75 and specificity of 0.89, and a REVEL score of 0.75, which gave a sensitivity of 0.55 and a specificity of 0.97 ^4^. Alternative thresholds were suggested by Pejaver *et al* ^2^ who evaluated the predictive power of the REVEL scores for pathogenic and benign variants in ClinVar to recommended that a score of 0.773 could be used as moderate and a score of 0.932 strong evidence for pathogenicity of a variant when assigning pathogenicity using the ACMG guidelines ^1^. The ability of REVEL to accurately predict pathogenicity of a variant with relatively high sensitivity and specificity has led to the tool being incorporated into gene-specific ACMG guidelines by Variant Curation Expert Panels (VCEPs). These include *RYR1* variants causing malignant hyperthermia susceptibility ^22^, *ITGA2B/ITGB3* variants causing Glanzmann thrombasthenia ^23^, and *MYOC* variants causing glaucoma ^24^ - the latter of which is likely caused by a GoF mechanism.

In a previous study SIFT and PolyPhen, two widely used *in silico* tools, were shown to perform less well at predicting the pathogenicity of GoF compared to LoF variants ^25^. This study exploited the fact that GoF and LoF variants in three genes (*ABCC8, KCNJ11* and *GCK*) cause opposing disease phenotypes (monogenic diabetes and congenital hyperinsulinism), creating a unique resource for evaluating different disease mechanisms within the same genes. In this study, we utilised this highly curated dataset to evaluate the performance of REVEL for predicting the pathogenicity of LoF and GoF variants within the same genes.

## Materials and methods

To evaluate the performance of REVEL on LoF and GoF variants we studied the curated set of 133 pathogenic variants from Flanagan *et al*. ^25^. We excluded two variants as one was a start-loss variant and the other was a multi-nucleotide variant, which REVEL is not designed to evaluate. This resulted in a set of 131 different pathogenic missense variants in the *ABCC8* (n = 47), *KCNJ11* (n = 56), and *GCK* (n = 28) genes (Supplementary Table 1). 66 variants were LoF while 65 were GoF. The authors of the REVEL paper ^4^ confirmed that the variants used in this study were not included in the training dataset for REVEL.

We downloaded the REVEL 1.3 dataset and looked up the REVEL scores for the 131 variants included in this study and evaluated the different thresholds for pathogenicity. This included REVEL scores of 0.5 and 0.75 as recommended by the authors of the tool ^4^. We also investigated the REVEL thresholds recommended by Pejaver *et al* ^2^ for using REVEL scores as different levels of evidence for pathogenicity (0.773 moderate and 0.932 strong).

Statistical significance was tested using Fisher’s Exact Test.

## Results

### Using author recommended thresholds REVEL correctly predicts pathogenicity of LoF and GoF variants

The authors of REVEL recommend potential thresholds for pathogenicity of REVEL scores of 0.5 or 0.75 depending on the context in which the tool was to be used – whether specificity or sensitivity was most important ^4^.

Using a 0.5 REVEL threshold for pathogenicity 65/66 (98%) LoF and 65/65 (100%) GoF variants were predicted as pathogenic. Using a 0.75 REVEL threshold for pathogenicity, 59/66 (89%) LoF and 57/65 (88%) GoF variants were predicted as pathogenic (Figure 1).

**Figure 1:**
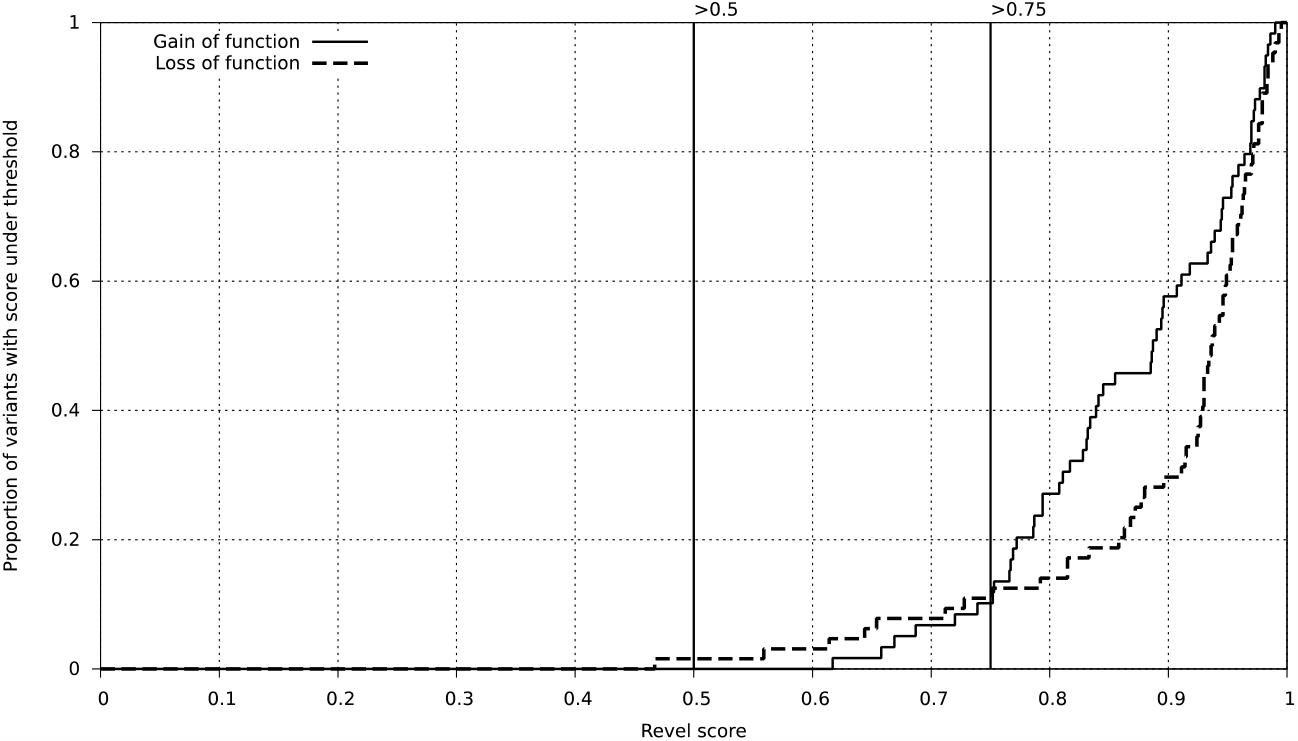
A high proportion of LoF and GoF variants meet author recommended thresholds for pathogenicity. A graph showing the cumulative frequency of loss of function (LoF) and gain of function (GoF) variants which meet that REVEL score threshold. The REVEL score thresholds of 0.5 and 0.75 are highlighted as they were given by the tool authors as potential thresholds for pathogenicity ^4^.

### REVEL scores for LoF variants are more likely to meet criteria for strong evidence for pathogenicity

The REVEL scores for 36/66 (55%) LoF variants meet the criteria for strong evidence (REVEL score of 0.932) as recommended by Pejaver *et al* ^2^ (Figure 2). In contrast only 23/65 (35%) GoF variants meet the criteria to use REVEL as strong evidence for pathogenicity (P=0.0352).

**Figure 2:**
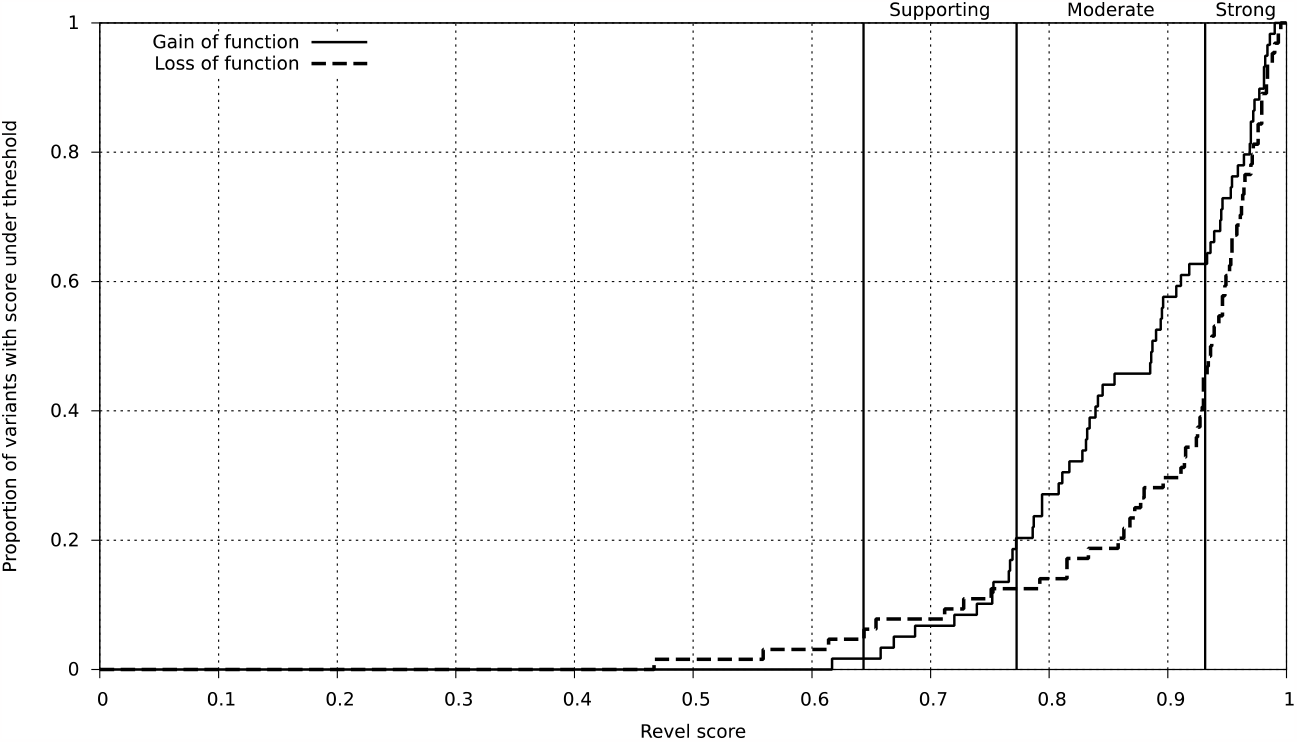
LoF variants are more likely to meet threshold for strong evidence for pathogenicity. A graph showing the cumulative frequency of loss of function (LoF) and gain of function (GoF) variants which meet that REVEL score threshold. The REVEL score thresholds for supporting, moderate and strong evidence are highlighted as recommended by Pejaver *et al*. ^2^.

Similarly, 58/66 (88%) of LoF variants meet at least the threshold for moderate evidence (REVEL score of 0.773) while 51/65 (78%) GoF variants meet that threshold, although this difference is not statistically significant (P=0.1677). 62/66 (94%) LoF and 63/65 (97%) GoF variants meet at least the criteria for supporting evidence (REVEL score of 0.644).

## Discussion

We used a dataset of 66 LoF and 65 GoF variants to assess the performance of the widely used meta-predictor REVEL for identifying pathogenic LoF and GoF variants. Using the REVEL score thresholds recommended by the authors of the tool (0.5 and 0.75) ^4^, REVEL performed similarly for LoF and GoF. However, when we then used the threshold recommended by Pejaver *et al* ^2^ as strong evidence of pathogenicity (REVEL score 0.932), a greater proportion of LoF than GoF variants met the criteria for strong evidence of pathogenicity.

There is not a clear pattern for the GoF variants that met the threshold for strong: they are split between the three genes studied and spread across protein domains. For example in *ABCC8*, using the protein domain classifications from De Franco *et al* ^26^, the variants that met the criteria for strong were split between the highly-conserved nucleotide-binding domain (n=3/6), transmembrane domain (n=2/6) and cytoplasmic domain (n=1/6). In comparison the variants which did not meet the criteria for strong were in the transmembrane (n=3/17), cytoplasmic (n=13/17) and extracellular (n=1/17) domains. This suggests that while protein domain may affect the REVEL score it is not deterministic of whether a variant will meet the threshold to be used as strong evidence for pathogenicity.

Our results suggest that GoF variants are less likely than LoF variants to get the very highest REVEL scores that would enable them to be used as strong evidence for pathogenicity. This is in keeping with the previous findings of Flanagan *et al*. ^25^ for SIFT and PolyPhen which found their predictive power was lower for these GoF variants. Since REVEL includes SIFT and PolyPhen2 scores as part of its algorithm, this may explain some of this difference in performance.

*In silico* predictors are not a substitute for expert judgement and should not be used in isolation but as part of an overall assessment of different strands of evidence as recommended by the ACMG guidelines ^1; 27^. Even if a REVEL score meets the threshold to use as strong evidence, further independent strands of evidence need to be provided for a variant to meet the ACMG criteria for pathogenicity. However, the ability to use the score from an *in silico* predictive tool as strong evidence of pathogenicity has important implications for variant classification. The ACMG guidelines state that two strong pieces of evidence are sufficient to declare a variant is pathogenic ^1^. A variant with a sufficiently high REVEL score would therefore only need one additional piece of strong evidence, such as *in vitro* functional evidence, to demonstrate pathogenicity. In contrast, if the *in silico* evidence can only be used as supporting then in addition to a strong piece of evidence you would also need two moderate (such as the variant being located in a well-established functional domain without benign variation) and a second supporting piece of evidence (such as the patient’s phenotype being highly specific for the disease), for example, to meet the threshold for pathogenicity. Some VCEPs have conservatively chosen to cap the use of REVEL scores for PP3 criteria to moderate ^22^ or supporting ^23^, which would mitigate the potential difference between the REVEL scores of GoF and LoF variants.

The finding that GoF variants are less likely than LoF variants to meet the score threshold to use as strong evidence of pathogenicity is important for diagnostic genetic testing of genes that cause disease via a GoF mechanism. This highlights a potential utility in developing bespoke REVEL thresholds for specific genetic conditions caused by GoF variants that would enable the scores to be used as moderate or strong evidence of pathogenicity. Indeed Pramparo *et al*. ^28^ calculated a bespoke REVEL cut off for pathogenic variants in *CYP27A1* causing cerebrotendinous xanthomatosis to study the prevalence and geographic distribution of the disease.

Variants in this study come from three genes where we have the expertise to confidently define whether they act via a LoF or GoF mechanism, since the two mechanisms cause the opposite phenotypes of monogenic diabetes or congenital hyperinsulinism. We did not include benign variants in the study as our aim was to evaluate the relative performance of REVEL on LoF and GoF variants rather than to assess the ability of the tool to accurately predict whether a variant was pathogenic or benign, which has already been established ^4; 21^. Whilst we expect our results to be widely applicable we recognise that our study was limited to three disease genes and we therefore recommend that further studies are performed on additional genes with known LoF and GoF mechanisms of pathogenicity in order to replicate our findings.

In conclusion, we found that REVEL correctly predicts a high proportion of both LoF and GoF variants as pathogenic based on the REVEL score thresholds recommended by the tool authors. However, GoF variants are less likely to receive the highest REVEL scores, which would preclude the score from being used as strong evidence of pathogenicity in some cases.

## Data Availability

The list of variants used in this study are included in Supplementary Table 1.

## Supplementary material

Supplementary Table 1 contains the list of variants used in this study.

A preprint has previously been published ^29^.

## Data availability

The list of variants used in this study are included in Supplementary Table 1.

## Acknowledgements

We would like to thank the authors of the REVEL paper, and specifically Joseph Rothstein, for confirming our variants did not form part of the REVEL training dataset.

## Author contributions

J.J.H, M.B.J and S.E.F curated the variants. M.N.W ran REVEL and generated the figures. T.W.L and M.N.W performed statistical analysis. T.W.L drafted the manuscript. All authors reviewed and approved the manuscript.

## References

1. Richards, S., Aziz, N., Bale, S., Bick, D., Das, S., Gastier-Foster, J., Grody, W.W., Hegde, M., Lyon, E., Spector, E., Voelkerding, K., and Rehm, H.L. (2015). Standards and guidelines for the interpretation of sequence variants: a joint consensus recommendation of the American College of Medical Genetics and Genomics and the Association for Molecular Pathology. Genet Med 17, 405–423.

2. Pejaver, V., Byrne, A.B., Feng, B.J., Pagel, K.A., Mooney, S.D., Karchin, R., O’Donnell-Luria, A., Harrison, S.M., Tavtigian, S.V., Greenblatt, M.S., Biesecker, L.G., Radivojac, P., and Brenner, S.E. (2022). Calibration of computational tools for missense variant pathogenicity classification and ClinGen recommendations for PP3/BP4 criteria. Am J Hum Genet 109, 2163–2177.

3. Rehm, H.L., Berg, J.S., Brooks, L.D., Bustamante, C.D., Evans, J.P., Landrum, M.J., Ledbetter, D.H., Maglott, D.R., Martin, C.L., Nussbaum, R.L., Plon, S.E., Ramos, E.M., Sherry, S.T., and Watson, M.S. (2015). ClinGen — The Clinical Genome Resource. New England Journal of Medicine 372, 2235–2242.

4. Ioannidis, N.M., Rothstein, J.H., Pejaver, V., Middha, S., McDonnell, S.K., Baheti, S., Musolf, A., Li, Q., Holzinger, E., Karyadi, D., Cannon-Albright, L.A., Teerlink, C.C., Stanford, J.L., Isaacs, W.B., Xu, J., Cooney, K.A., Lange, E.M., Schleutker, J., Carpten, J.D., Powell, I.J., Cussenot, O., Cancel-Tassin, G., Giles, G.G., MacInnis, R.J., Maier, C., Hsieh, C.L., Wiklund, F., Catalona, W.J., Foulkes, W.D., Mandal, D., Eeles, R.A., Kote-Jarai, Z., Bustamante, C.D., Schaid, D.J., Hastie, T., Ostrander, E.A., Bailey-Wilson, J.E., Radivojac, P., Thibodeau, S.N., Whittemore, A.S., and Sieh, W. (2016). REVEL: An Ensemble Method for Predicting the Pathogenicity of Rare Missense Variants. Am J Hum Genet 99, 877–885.

5. Pejaver, V., Urresti, J., Lugo-Martinez, J., Pagel, K.A., Lin, G.N., Nam, H.-J., Mort, M., Cooper, D.N., Sebat, J., Iakoucheva, L.M., Mooney, S.D., and Radivojac, P. (2020). Inferring the molecular and phenotypic impact of amino acid variants with MutPred2. Nature Communications 11, 5918.

6. Shihab, H.A., Gough, J., Cooper, D.N., Stenson, P.D., Barker, G.L., Edwards, K.J., Day, I.N., and Gaunt, T.R. (2013). Predicting the functional, molecular, and phenotypic consequences of amino acid substitutions using hidden Markov models. Hum Mutat 34, 57–65.

7. Carter, H., Douville, C., Stenson, P.D., Cooper, D.N., and Karchin, R. (2013). Identifying Mendelian disease genes with the variant effect scoring tool. BMC Genomics 14 Suppl 3, S3.

8. Adzhubei, I.A., Schmidt, S., Peshkin, L., Ramensky, V.E., Gerasimova, A., Bork, P., Kondrashov, A.S., and Sunyaev, S.R. (2010). A method and server for predicting damaging missense mutations. Nature Methods 7, 248–249.

9. Ng, P.C., and Henikoff, S. (2003). SIFT: Predicting amino acid changes that affect protein function. Nucleic Acids Res 31, 3812–3814.

10. Choi, Y., and Chan, A.P. (2015). PROVEAN web server: a tool to predict the functional effect of amino acid substitutions and indels. Bioinformatics 31, 2745–2747.

11. Reva, B., Antipin, Y., and Sander, C. (2011). Predicting the functional impact of protein mutations: application to cancer genomics. Nucleic Acids Res 39, e118.

12. Schwarz, J.M., Rödelsperger, C., Schuelke, M., and Seelow, D. (2010). MutationTaster evaluates disease-causing potential of sequence alterations. Nature Methods 7, 575–576.

13. Chun, S., and Fay, J.C. (2009). Identification of deleterious mutations within three human genomes. Genome Res 19, 1553–1561.

14. Davydov, E.V., Goode, D.L., Sirota, M., Cooper, G.M., Sidow, A., and Batzoglou, S. (2010). Identifying a high fraction of the human genome to be under selective constraint using GERP++. PLoS Comput Biol 6, e1001025.

15. Garber, M., Guttman, M., Clamp, M., Zody, M.C., Friedman, N., and Xie, X. (2009). Identifying novel constrained elements by exploiting biased substitution patterns. Bioinformatics 25, i54–62.

16. Pollard, K.S., Hubisz, M.J., Rosenbloom, K.R., and Siepel, A. (2010). Detection of nonneutral substitution rates on mammalian phylogenies. Genome Res 20, 110–121.

17. Siepel, A., Bejerano, G., Pedersen, J.S., Hinrichs, A.S., Hou, M., Rosenbloom, K., Clawson, H., Spieth, J., Hillier, L.W., Richards, S., Weinstock, G.M., Wilson, R.K., Gibbs, R.A., Kent, W.J., Miller, W., and Haussler, D. (2005). Evolutionarily conserved elements in vertebrate, insect, worm, and yeast genomes. Genome Res 15, 1034–1050.

18. Toratani, K., Watanabe, M., Kanda, J., Oka, T., Hyuga, M., Arai, Y., Iwasaki, M., Sakurada, M., Nannya, Y., Ogawa, S., Yamada, T., and Takaori-Kondo, A. (2023). Unrelated hematopoietic stem cell transplantation for familial platelet disorder/acute myeloid leukemia with germline RUNX1 mutations. Int J Hematol.

19. Schuurmans, I.M.E., Dimitrov, B., Schröter, J., Ribes, A., de la Fuente, R.P., Zamora, B., van Karnebeek, C.D.M., Kölker, S., and Garanto, A. (2023). Exploring genotype-phenotype correlations in glutaric aciduria type 1. J Inherit Metab Dis.

20. Kingdom, R., Tuke, M., Wood, A., Beaumont, R.N., Frayling, T.M., Weedon, M.N., and Wright, C.F. (2022). Rare genetic variants in genes and loci linked to dominant monogenic developmental disorders cause milder related phenotypes in the general population. Am J Hum Genet 109, 1308–1316.

21. Gunning, A.C., Fryer, V., Fasham, J., Crosby, A.H., Ellard, S., Baple, E.L., and Wright, C.F. (2021). Assessing performance of pathogenicity predictors using clinically relevant variant datasets. Journal of medical genetics 58, 547–555.

22. Johnston, J.J., Dirksen, R.T., Girard, T., Gonsalves, S.G., Hopkins, P.M., Riazi, S., Saddic, L.A., Sambuughin, N., Saxena, R., Stowell, K., Weber, J., Rosenberg, H., and Biesecker, L.G. (2021). Variant curation expert panel recommendations for RYR1 pathogenicity classifications in malignant hyperthermia susceptibility. Genet Med 23, 1288–1295.

23. Ross, J.E., Zhang, B.M., Lee, K., Mohan, S., Branchford, B.R., Bray, P., Dugan, S.N., Freson, K., Heller, P.G., Kahr, W.H.A., Lambert, M.P., Luchtman-Jones, L., Luo, M., Perez Botero, J., Rondina, M.T., Ryan, G., Westbury, S., Bergmeier, W., and Di Paola, J. (2021). Specifications of the variant curation guidelines for ITGA2B/ITGB3: ClinGen Platelet Disorder Variant Curation Panel. Blood Adv 5, 414–431.

24. Burdon, K.P., Graham, P., Hadler, J., Hulleman, J.D., Pasutto, F., Boese, E.A., Craig, J.E., Fingert, J.H., Hewitt, A.W., Siggs, O.M., Whisenhunt, K., Young, T.L., Mackey, D.A., Dubowsky, A., and Souzeau, E. (2022). Specifications of the ACMG/AMP variant curation guidelines for myocilin: Recommendations from the clingen glaucoma expert panel. Hum Mutat 43, 2170–2186.

25. Flanagan, S.E., Patch, A.M., and Ellard, S. (2010). Using SIFT and PolyPhen to predict loss-of-function and gain-of-function mutations. Genet Test Mol Biomarkers 14, 533–537.

26. De Franco, E., Saint-Martin, C., Brusgaard, K., Knight Johnson, A.E., Aguilar-Bryan, L., Bowman, P., Arnoux, J.B., Larsen, A.R., Sanyoura, M., Greeley, S.A.W., Calzada-León, R., Harman, B., Houghton, J.A.L., Nishimura-Meguro, E., Laver, T.W., Ellard, S., Del Gaudio, D., Christesen, H.T., Bellanné-Chantelot, C., and Flanagan, S.E. (2020). Update of variants identified in the pancreatic β-cell K(ATP) channel genes KCNJ11 and ABCC8 in individuals with congenital hyperinsulinism and diabetes. Hum Mutat 41, 884–905.

27. Ellard, S., Colclough, K., Patel, K.A., and Hattersley, A.T. (2020). Prediction algorithms: pitfalls in interpreting genetic variants of autosomal dominant monogenic diabetes. J Clin Invest 130, 14–16.

28. Pramparo, T., Steiner, R.D., Rodems, S., and Jenkinson, C. (2023). Allelic prevalence and geographic distribution of cerebrotendinous xanthomatosis. Orphanet J Rare Dis 18, 13.

29. Hopkins, J.J., Wakeling, M.N., Johnson, M.B., Flanagan, S.E., and Laver, T.W. (2023). REVEL is better at predicting pathogenicity of loss-of-function than gain-of-function variants. medRxiv, 2023.2006.2006.23290963.

