## Supplementary Table 1 for "REVEL is better at predicting pathogenicity of loss-of-function than gain-of-function variants"

**Supplementary Table 1: Variants used to evaluate the performance of REVEL on loss of function (LoF) and gain of function (GoF) variants.** Diabetes includes neonatal diabetes and maturity onset diabetes of the young (MODY). HI; hyperinsulinism. Nomenclature is given according to the following transcripts: *ABCC8* NM_001287174.1, *GCK* NM_000162.3 and *KCNJ11* NM_000525.4.

| **Gene** | **Protein change** | **Nucleotide change** | **Genomic location (GRCh37)** | **REVEL score** | **GoF or LoF** | **Phenotype** |
| --- | --- | --- | --- | --- | --- | --- |
| *ABCC8* | p.(Val86Ala) | c.257T>C | 11:17496466A>G | 0.794 | GoF | Diabetes |
| *ABCC8* | p.(Val86Gly) | c.257T>G | 11:17496466A>C | 0.97 | GoF | Diabetes |
| *ABCC8* | p.(Phe132Leu) | c.394T>C | 11:17491666A>G | 0.89 | GoF | Diabetes |
| *ABCC8* | p.(Phe132Val) | c.394T>G | 11:17491666A>C | 0.808 | GoF | Diabetes |
| *ABCC8* | p.(Glu208Lys) | c.622G>A | 11:17483330C>T | 0.669 | GoF | Diabetes |
| *ABCC8* | p.(Asp209Glu) | c.627C>A | 11:17483325G>T | 0.739 | GoF | Diabetes |
| *ABCC8* | p.(Gln211Lys) | c.631C>A | 11:17483321G>T | 0.617 | GoF | Diabetes |
| *ABCC8* | p.(Leu213Arg) | c.638T>G | 11:17483314A>C | 0.72 | GoF | Diabetes |
| *ABCC8* | p.(Leu225Pro) | c.674T>C | 11:17483278A>G | 0.769 | GoF | Diabetes |
| *ABCC8* | p.(Thr229Ile) | c.686C>T | 11:17483266G>A | 0.894 | GoF | Diabetes |
| *ABCC8* | p.(Val324Met) | c.970G>A | 11:17482076C>T | 0.895 | GoF | Diabetes |
| *ABCC8* | p.(Leu451Pro) | c.1352T>C | 11:17464840A>G | 0.936 | GoF | Diabetes |
| *ABCC8* | p.(Leu582Val) | c.1744C>G | 11:17452434G>C | 0.839 | GoF | Diabetes |
| *ABCC8* | p.(Arg826Trp) | c.2476C>T | 11:17434943G>A | 0.855 | GoF | Diabetes |
| *ABCC8* | p.(His1024Tyr) | c.3070C>T | 11:17428530G>A | 0.687 | GoF | Diabetes |
| *ABCC8* | p.(Asn1123Asp) | c.3367A>G | 11:17427076T>C | 0.896 | GoF | Diabetes |
| *ABCC8* | p.(Arg1183Trp) | c.3547C>T | 11:17426072G>A | 0.885 | GoF | Diabetes |
| *ABCC8* | p.(Arg1183Gln) | c.3548G>A | 11:17426071C>T | 0.767 | GoF | Diabetes |
| *ABCC8* | p.(Arg1314His) | c.3941G>A | 11:17418790C>T | 0.933 | GoF | Diabetes |
| *ABCC8* | p.(Arg1380Cys) | c.4138C>T | 11:17417462G>A | 0.953 | GoF | Diabetes |
| *ABCC8* | p.(Arg1380His) | c.4139G>A | 11:17417461C>T | 0.964 | GoF | Diabetes |
| *ABCC8* | p.(Arg1380Leu) | c.4139G>T | 11:17417461C>A | 0.959 | GoF | Diabetes |
| *ABCC8* | p.(Ile1425Val) | c.4273A>G | 11:17417194T>C | 0.772 | GoF | Diabetes |
| *ABCC8* | p.(Gly7Arg) | c.19G>C | 11:17498305C>G | 0.954 | LoF | HI |
| *ABCC8* | p.(Val21Asp) | c.62T>A | 11:17498262A>T | 0.863 | LoF | HI |
| *ABCC8* | p.(Asn24Lys) | c.72C>A | 11:17498252G>T | 0.858 | LoF | HI |
| *ABCC8* | p.(Arg74Trp) | c.220C>T | 11:17496503G>A | 0.976 | LoF | HI |
| *ABCC8* | p.(Gly111Trp) | c.331G>T | 11:17491729C>A | 0.636 | LoF | HI |
| *ABCC8* | p.(His125Gln) | c.375C>G | 11:17491685G>C | 0.888 | LoF | HI |
| *ABCC8* | p.(Val187Asp) | c.560T>A | 11:17485004A>T | 0.815 | LoF | HI |
| *ABCC8* | p.(Gly228Asp) | c.683G>A | 11:17483269C>T | 0.614 | LoF | HI |
| *ABCC8* | p.(Asp310Asn) | c.928G>A | 11:17482118C>T | 0.833 | LoF | HI |
| *ABCC8* | p.(Arg370Gly) | c.1108A>G | 11:17474734T>C | 0.644 | LoF | HI |
| *ABCC8* | p.(Arg370Ser) | c.1110G>C | 11:17474732C>G | 0.559 | LoF | HI |
| *ABCC8* | p.(Arg495Gln) | c.1484G>A | 11:17464413C>T | 0.872 | LoF | HI |
| *ABCC8* | p.(Arg1215Trp) | c.3643C>T | 11:17424218G>A | 0.964 | LoF | HI |
| *ABCC8* | p.(Arg1353His) | c.4058G>A | 11:17418527C>T | 0.934 | LoF | HI |
| *ABCC8* | p.(Lys1374Arg) | c.4121A>G | 11:17418464T>C | 0.868 | LoF | HI |
| *ABCC8* | p.(Ser1386Pro) | c.4156T>C | 11:17417444A>G | 0.979 | LoF | HI |
| *ABCC8* | p.(Gly1478Val) | c.4433G>T | 11:17415928C>A | 0.88 | LoF | HI |
| *ABCC8* | p.(Gly1479Arg) | c.4435G>A | 11:17415926C>T | 0.939 | LoF | HI |
| *ABCC8* | p.(Arg1494Trp) | c.4480C>T | 11:17415881G>A | 0.93 | LoF | HI |
| *ABCC8* | p.(Arg1494Gln) | c.4481G>A | 11:17415880C>T | 0.979 | LoF | HI |
| *ABCC8* | p.(Glu1507Lys) | c.4519G>A | 11:17415842C>T | 0.963 | LoF | HI |
| *ABCC8* | p.(Ile1512Thr) | c.4535T>C | 11:17415826A>G | 0.927 | LoF | HI |
| *ABCC8* | p.(Arg1539Gln) | c.4616G>A | 11:17414671C>T | 0.877 | LoF | HI |
| *ABCC8* | p.(Leu1544Pro) | c.4631T>C | 11:17414656A>G | 0.915 | LoF | HI |
| *GCK* | p.(Ser64Tyr) | c.191C>A | 7:44192917G>T | 0.984 | GoF | HI |
| *GCK* | p.(Thr65Ile) | c.194C>T | 7:44192914G>A | 0.82 | GoF | HI |
| *GCK* | p.(Gly68Val) | c.203G>T | 7:44192905C>A | 0.993 | GoF | HI |
| *GCK* | p.(Trp99Arg) | c.295T>C | 7:44191938A>G | 0.753 | GoF | HI |
| *GCK* | p.(Tyr214Cys) | c.641A>G | 7:44189397T>C | 0.706 | GoF | HI |
| *GCK* | p.(Val452Leu) | c.1354G>C | 7:44184779C>G | 0.601 | GoF | HI |
| *GCK* | p.(Val455Met) | c.1363G>A | 7:44184770C>T | 0.853 | GoF | HI |
| *GCK* | p.(Ala456Val) | c.1367C>T | 7:44184766G>A | 0.883 | GoF | HI |
| *GCK* | p.(Gly44Ser) | c.130G>A | 7:44192978C>T | 0.962 | LoF | Diabetes |
| *GCK* | p.(Gly44Asp) | c.131G>A | 7:44192977C>T | 0.995 | LoF | Diabetes |
| *GCK* | p.(Tyr108His) | c.322T>C | 7:44191911A>G | 0.949 | LoF | Diabetes |
| *GCK* | p.(Cys129Tyr) | c.386G>A | 7:44190652C>T | 0.97 | LoF | Diabetes |
| *GCK* | p.(Phe150Ser) | c.449T>C | 7:44190589A>G | 0.983 | LoF | Diabetes |
| *GCK* | p.(Asp160Asn) | c.478G>A | 7:44190560C>T | 0.712 | LoF | Diabetes |
| *GCK* | p.(Asn180Lys) | c.540T>G | 7:44189607A>C | 0.751 | LoF | Diabetes |
| *GCK* | p.(Val182Met) | c.544G>A | 7:44189603C>T | 0.937 | LoF | Diabetes |
| *GCK* | p.(Ala188Thr) | c.562G>A | 7:44189585C>T | 0.933 | LoF | Diabetes |
| *GCK* | p.(Arg191Trp) | c.571C>T | 7:44189576G>A | 0.936 | LoF | Diabetes |
| *GCK* | p.(Ala201Ser) | c.601G>T | 7:44189437C>A | 0.929 | LoF | Diabetes |
| *GCK* | p.(Val203Ala) | c.608T>C | 7:44189430A>G | 0.946 | LoF | Diabetes |
| *GCK* | p.(Gly223Ser) | c.667G>A | 7:44189371C>T | 0.943 | LoF | Diabetes |
| *GCK* | p.(Gly261Arg) | c.781G>A | 7:44187331C>T | 0.914 | LoF | Diabetes |
| *GCK* | p.(Glu265Lys) | c.793G>A | 7:44187319C>T | 0.728 | LoF | Diabetes |
| *GCK* | p.(Asp278Glu) | c.834C>A | 7:44187278G>T | 0.896 | LoF | Diabetes |
| *GCK* | p.(Ser340Ile) | c.1019G>T | 7:44186062C>A | 0.792 | LoF | Diabetes |
| *GCK* | p.(Met381Thr) | c.1142T>C | 7:44185207A>G | 0.93 | LoF | Diabetes |
| *GCK* | p.(Arg392Cys) | c.1174C>T | 7:44185175G>A | 0.911 | LoF | Diabetes |
| *GCK* | p.(Arg397Leu) | c.1190G>T | 7:44185159C>A | 0.925 | LoF | Diabetes |
| *KCNJ11* | p.(Ser3Cys) | c.8C>G | 11:17409631G>C | 0.658 | GoF | Diabetes |
| *KCNJ11* | p.(His46Tyr) | c.136C>T | 11:17409503G>A | 0.845 | GoF | Diabetes |
| *KCNJ11* | p.(Arg50Gln) | c.149G>A | 11:17409490C>T | 0.831 | GoF | Diabetes |
| *KCNJ11* | p.(Arg50Pro) | c.149G>C | 11:17409490C>G | 0.887 | GoF | Diabetes |
| *KCNJ11* | p.(Glu51Ala) | c.152A>C | 11:17409487T>G | 0.907 | GoF | Diabetes |
| *KCNJ11* | p.(Gln52Arg) | c.155A>G | 11:17409484T>C | 0.811 | GoF | Diabetes |
| *KCNJ11* | p.(Gly53Ser) | c.157G>A | 11:17409482C>T | 0.794 | GoF | Diabetes |
| *KCNJ11* | p.(Gly53Arg) | c.157G>C | 11:17409482C>G | 0.817 | GoF | Diabetes |
| *KCNJ11* | p.(Gly53Asp) | c.158G>A | 11:17409481C>T | 0.841 | GoF | Diabetes |
| *KCNJ11* | p.(Val59Met) | c.175G>A | 11:17409464C>T | 0.766 | GoF | Diabetes |
| *KCNJ11* | p.(Val59Gly) | c.176T>G | 11:17409463A>C | 0.939 | GoF | Diabetes |
| *KCNJ11* | p.(Leu164Pro) | c.491T>C | 11:17409148A>G | 0.973 | GoF | Diabetes |
| *KCNJ11* | p.(Cys166Tyr) | c.497G>A | 11:17409142C>T | 0.954 | GoF | Diabetes |
| *KCNJ11* | p.(Cys166Phe) | c.497G>T | 11:17409142C>A | 0.944 | GoF | Diabetes |
| *KCNJ11* | p.(Ile167Leu) | c.499A>C | 11:17409140T>G | 0.828 | GoF | Diabetes |
| *KCNJ11* | p.(Lys170Thr) | c.509A>C | 11:17409130T>G | 0.99 | GoF | Diabetes |
| *KCNJ11* | p.(Lys170Arg) | c.509A>G | 11:17409130T>C | 0.946 | GoF | Diabetes |
| *KCNJ11* | p.(Lys170Asn) | c.510G>C | 11:17409129C>G | 0.886 | GoF | Diabetes |
| *KCNJ11* | p.(Glu179Ala) | c.536A>C | 11:17409103T>G | 0.945 | GoF | Diabetes |
| *KCNJ11* | p.(Ile182Val) | c.544A>G | 11:17409095T>C | 0.786 | GoF | Diabetes |
| *KCNJ11* | p.(Arg201Ser) | c.601C>A | 11:17409038G>T | 0.969 | GoF | Diabetes |
| *KCNJ11* | p.(Arg201Cys) | c.601C>T | 11:17409038G>A | 0.977 | GoF | Diabetes |
| *KCNJ11* | p.(Arg201His) | c.602G>A | 11:17409037C>T | 0.982 | GoF | Diabetes |
| *KCNJ11* | p.(Arg201Leu) | c.602G>T | 11:17409037C>A | 0.986 | GoF | Diabetes |
| *KCNJ11* | p.(Glu227Lys) | c.679G>A | 11:17408960C>T | 0.972 | GoF | Diabetes |
| *KCNJ11* | p.(Glu229Lys) | c.685G>A | 11:17408954C>T | 0.981 | GoF | Diabetes |
| *KCNJ11* | p.(Leu233Phe) | c.697C>T | 11:17408942G>A | 0.752 | GoF | Diabetes |
| *KCNJ11* | p.(Val252Met) | c.754G>A | 11:17408885C>T | 0.918 | GoF | Diabetes |
| *KCNJ11* | p.(Val252Gly) | c.755T>G | 11:17408884A>C | 0.97 | GoF | Diabetes |
| *KCNJ11* | p.(Thr293Asn) | c.878C>A | 11:17408761G>T | 0.832 | GoF | Diabetes |
| *KCNJ11* | p.(Ile296Leu) | c.886A>C | 11:17408753T>G | 0.834 | GoF | Diabetes |
| *KCNJ11* | p.(Glu322Lys) | c.964G>A | 11:17408675C>T | 0.911 | GoF | Diabetes |
| *KCNJ11* | p.(Tyr330Ser) | c.989A>C | 11:17408650T>G | 0.981 | GoF | Diabetes |
| *KCNJ11* | p.(Gly334Asp) | c.1001G>A | 11:17408638C>T | 0.787 | GoF | Diabetes |
| *KCNJ11* | p.(Arg34His) | c.101G>A | 11:17409538C>T | 0.965 | LoF | HI |
| *KCNJ11* | p.(Gly40Asp) | c.119G>A | 11:17409520C>T | 0.988 | LoF | HI |
| *KCNJ11* | p.(Phe55Leu) | c.165C>A | 11:17409474G>T | 0.924 | LoF | HI |
| *KCNJ11* | p.(Thr62Met) | c.185C>T | 11:17409454G>A | 0.952 | LoF | HI |
| *KCNJ11* | p.(Lys67Asn) | c.201G>C | 11:17409438C>G | 0.815 | LoF | HI |
| *KCNJ11* | p.(Trp91Arg) | c.271T>C | 11:17409368A>G | 0.953 | LoF | HI |
| *KCNJ11* | p.(Ala101Asp) | c.302C>A | 11:17409337G>T | 0.467 | LoF | HI |
| *KCNJ11* | p.(Ser116Pro) | c.346T>C | 11:17409293A>G | 0.93 | LoF | HI |
| *KCNJ11* | p.(Gly134Ala) | c.401G>C | 11:17409238C>G | 0.976 | LoF | HI |
| *KCNJ11* | p.(Arg136Leu) | c.407G>T | 11:17409232C>A | 0.984 | LoF | HI |
| *KCNJ11* | p.(Gly156Arg) | c.466G>A | 11:17409173C>T | 0.99 | LoF | HI |
| *KCNJ11* | p.(Asp204Glu) | c.612C>A | 11:17409027G>T | 0.948 | LoF | HI |
| *KCNJ11* | p.(Ala213Thr) | c.637G>A | 11:17409002C>T | 0.954 | LoF | HI |
| *KCNJ11* | p.(Pro254Leu) | c.761C>T | 11:17408878G>A | 0.971 | LoF | HI |
| *KCNJ11* | p.(His259Arg) | c.776A>G | 11:17408863T>C | 0.993 | LoF | HI |
| *KCNJ11* | p.(Pro266Leu) | c.797C>T | 11:17408842G>A | 0.979 | LoF | HI |
| *KCNJ11* | p.(Glu282Lys) | c.844G>A | 11:17408795C>T | 0.961 | LoF | HI |
| *KCNJ11* | p.(Thr294Met) | c.881C>T | 11:17408758G>A | 0.958 | LoF | HI |
| *KCNJ11* | p.(Arg301Gly) | c.901C>G | 11:17408738G>C | 0.946 | LoF | HI |
| *KCNJ11* | p.(Arg301Cys) | c.901C>T | 11:17408738G>A | 0.94 | LoF | HI |
| *KCNJ11* | p.(Arg301His) | c.902G>A | 11:17408737C>T | 0.972 | LoF | HI |
| *KCNJ11* | p.(Arg301Pro) | c.902G>C | 11:17408737C>G | 0.984 | LoF | HI |
